# Similar HbA1c, Similar BMI, Different disease: The Adipo-B Index Reveals Hidden Metabolic Heterogeneity in Newly Diagnosed Japanese Subjects with Type 2 Diabetes

**DOI:** 10.64898/2026.05.31.26354545

**Authors:** Eiji Kutoh, Alexandra N. Kuto

**Affiliations:** Biomedical Center, Tokyo, Japan; Division of Diabetes, Department of Internal Medicine, Kumagaya Surgery Hospital, Kumagaya, Japan; Division of Diabetes and Endocrinology, Department of Internal Medicine, Hamamatsu South Hospital, Hamamatsu, Japan

## Abstract

**Objective:** Patients and physicians frequently focus on HbA1c and weight alone. We hypothesized that individuals with similar HbA1c and BMI may present markedly distinct metabolic backgrounds. We investigated whether the adipo-B index- composite of adipose insulin resistance (adipo-IR) and beta-cell function (HOMA-B)-can uncover hidden heterogeneity in this clinically “homogeneous” population.

**Methods:** A total of 399 newly diagnosed, drug-naïve Japanese subjects with T2DM were analyzed. Histograms of HbA1c and BMI demonstrated peak distributions within HbA1c 8-10% and BMI 24-26 kg/m^2^. Based on these distributions, a clinically homogeneous subgroup was defined to minimize confounding by glycemic severity and adiposity. Metabolic parameters including FBG, insulin, FFA, HOMA-R, HOMA-B, adipo-IR, adipo-B, T-C, TG, HDL-C and non-HDL-C were analyzed. Simple regression, multivariable linear regression, and subgroup stratification analyses were performed.

**Results:** Despite comparable HbA1c and BMI by design, adipo-B stratification revealed significant differences in HOMA-B, FFA, non-HDL-C, and TG, whereas HOMA-R stratification identified only higher insulin and adipo-IR without differences in lipids or HOMA-B. Thus, adipo-B-but not HOMA-R-identified a lipotoxic, beta-cell-stressed phenotype invisible to conventional markers. Simple regression showed significant positive correlations between adipo-B and HbA1c, FBG, FFA, T-C, TG, and non-HDL-C, and negative correlations with insulin and HOMA-B. Multivariable linear regression confirmed that adipo-B was independently associated with non-HDL cholesterol, TG, and FFA after adjustment for HbA1c and BMI.

**Conclusion:** Even among patients with identical HbA1c and BMI, the adipo-B index uncovers clinically relevant metabolic heterogeneity, supporting its role as a functional marker of the adipose-pancreas axis and a potential tool for precision phenotyping in early T2DM.

## Introduction

T2DM is characterized by substantial metabolic heterogeneity involving variable degrees of insulin resistance, beta-cell dysfunction, adipose tissue dysregulation, and lipid abnormalities (1, 2, 3). However, in routine clinical practice, disease severity is primarily evaluated with HbA1c and body mass index (BMI). Although these parameters provide important information regarding glycemic control and obesity status, they may insufficiently reflect the complexity of the underlying metabolic phenotype. In other words, metabolic health is not determined solely by glycemia or body weight. This limitation may be particularly relevant in Japanese subjects with T2DM, who frequently develop diabetes at relatively modest BMI levels and often exhibit impaired beta-cell function disproportionate to obesity severity (4, 5). Consequently, individuals with similar HbA1c and BMI may still possess markedly different metabolic backgrounds and cardiovascular risk profiles.

Recently, adipose tissue-related metabolic indices have attracted attention as potential integrative markers of systemic metabolic dysfunction. Adipo-IR, calculated from fasting free fatty acid (FFA) and insulin levels, has been proposed as a surrogate marker of adipose tissue insulin resistance (6). As proposed in our previous work, adipose tissue insulin resistance (adipo-IR) and pancreatic beta-cell function (HOMA-B) could interact to determine systemic metabolic status (7). In this work, it is proposed that adipo-B index integrates these two axes by normalizing adipose-derived lipotoxic load (adipo-IR) against beta-cell compensatory capacity (HOMA-B), providing a functional measure of metabolic stress (7).

In the current work, a simple but clinically important question was asked: Do patients with the similar HbA1c and BMI truly have the same disease? Using a relatively large cohort of treatment-naïve subjects with T2DM, we isolated individuals with nearly identical HbA1c and BMI and examined whether adipo-B could differentiate their metabolic backgrounds. The present study investigated whether adipo-B could identify hidden metabolic heterogeneity among newly diagnosed Japanese subjects with T2DM who exhibited similar HbA1c and BMI values. To minimize confounding by glycemic severity and adiposity, we focused on a clinically homogeneous subgroup corresponding to the peak frequency distribution of HbA1c and BMI.

## Subjects and Methods

### Study Population

We analyzed 399 newly diagnosed, drug-naïve subjects with T2DM enrolled under the same criteria described in our previous study (8). All subjects met Japan Diabetes Society diagnostic criteria (9) and had no major comorbidities.

### Selection of a Homogeneous (Glycemia-Weight-Matched) Subset

To examine metabolic heterogeneity independent of glycemic severity and body size, we defined a “clinically homogeneous” subset as subjects with HbA1c within 8-10% and BMI within 24-26 kg/m^2^, based on data-driven peak distributions observed in the full cohort. This approach ensured that subsequent analyses were conducted within a narrow range of glycemia and adiposity, minimizing confounding by these structural markers. A total of 30 subjects met these criteria and were included in further analyses. Within this subset, participants were divided into two groups (low and high adipo-B) using median splits. This allowed evaluation of whether an integrated metabolic index (adipo-B) or an isolated insulin resistance index (HOMA-R) could differentiate metabolic phenotypes under conditions of matched HbA1c and BMI.

## Biochemical Measurements

Fasting blood samples were used to calculate the following indices (6, 7, 10).

HOMA-R=(insulin × FBG)/405

adipo-IR=FFA×insulin

HOMA-B=(insulin x× 360)/(FBG-63)

adipo-B= adipo-IR/HOMA-B

## Statistical Analysis

Group comparisons were performed using unpaired t-tests or Mann-Whitney tests as appropriate. Simple regression analysis was used to identify correlations between adipo-B and other metabolic parameters. Multivariable linear regression analysis was then performed to determine whether adipo-B was independently associated with the metabolic parameters identified in simple regression, after adjustment for HbA1c and BMI. For subgroup analyses, subjects were divided into low and high adipo-B groups using a median split. Statistical significance was set at p<0.05; p-values between 0.05 and 0.1 were considered to represent a trend toward significance (11). All analyses were performed using PAST software (University of Oslo; https://www.nhm.uio.no/english/research/resources/past/).

## Results

### Distribution of HbA1c and BMI in Newly Diagnosed Japanese Subjects with T2DM

Among 399 newly diagnosed, drug-naïve subjects with T2DM, the distributions of HbA1c and BMI demonstrated clear central peaks (Fig. 1A and 1B). The HbA1c histogram showed the highest frequency within the 8-10% range, whereas the BMI histogram peaked within 24-26 kg/m^2^. Based on these distributions, a clinically homogeneous subgroup was defined as subjects with HbA1c 8-10% and BMI 24-26 kg/m^2^. A total of 30 subjects met these criteria and were included in subsequent subgroup analyses.

**Figure 1.**
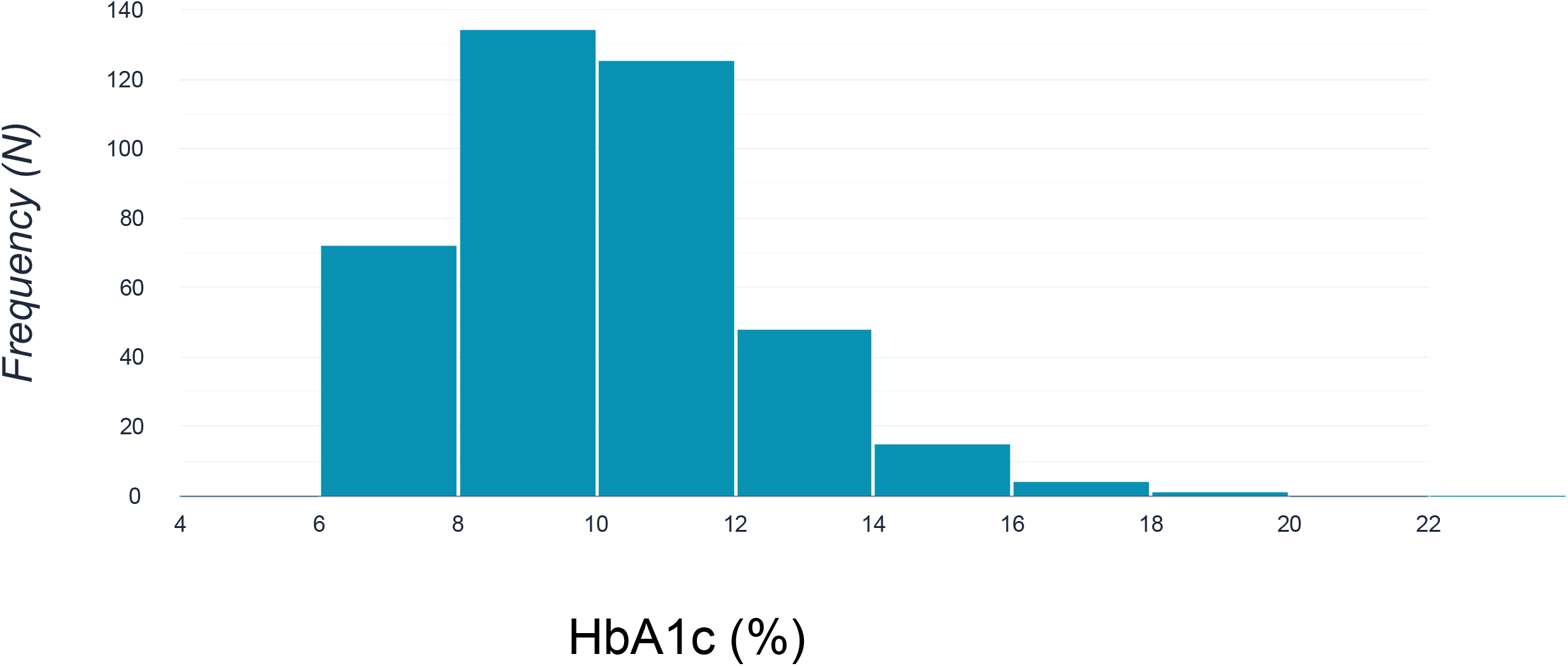
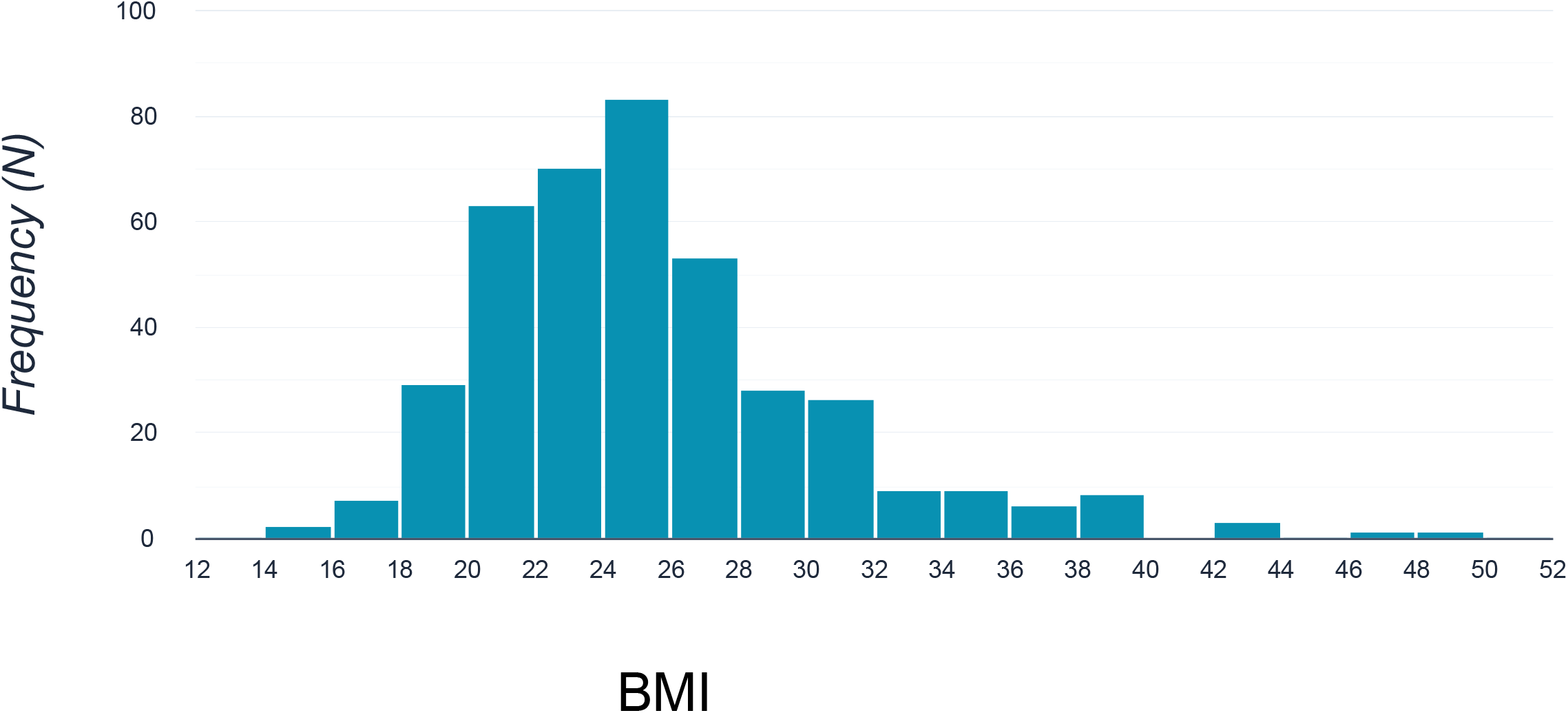
Frequency distributions of HbA1c and BMI in newly diagnosed, drug-naïve type 2 diabetes (N = 399). A) Histogram of HbA1c values in the full cohort. B) Histogram of BMI values in the same cohort.

### Correlations Between adipo-B and Metabolic Parameters

To investigate the relationship between adipo-B and metabolic parameters, simple regression analyses were performed (Table 2). Significant positive correlations were observed between adipo-B and HbA1c, FBG, FFA, T-C, TG, and non-HDL-C. In contrast, significant negative correlations were observed between adipo-B and insulin as well as HOMA-B.

**Table 1.** Characteristics of the selected subjects (HbA1c 8-10%, BMI 24-26 kg/m^2^) These subjects were selected from the peak area of the histogram (Fig. 1A and 1B).

|  |  |
| --- | --- |
| age | 56.31±10.65 |
| HbA1c (%) | 8.81±0.44 |
| BMI | 24.64±0.72 |
| adipo-B | 0.276±0.162 |
| adipo-IR | 4.96±2.43 |
| FBG (mg/dl) | 181.2±49.1 |
| insulin (μU/ml) | 6.29±2.73 |
| FFA (mEq/L) | 0.855±0.455 |
| HOMA-R | 2.81±1.42 |
| HOMA-B | 22.66±16.33 |
| T-C (mg/dl) | 223.8±52.6 |
| TG (mg/dl) | 243.2±247.7 |
| HDL-C (mg/dl) | 49.4±10.3 |
| non-HDL-C (mg/dl) | 174.3±51.7 |
| UA (mg/dl) | 5.22±1.16 |

**Table 2.**
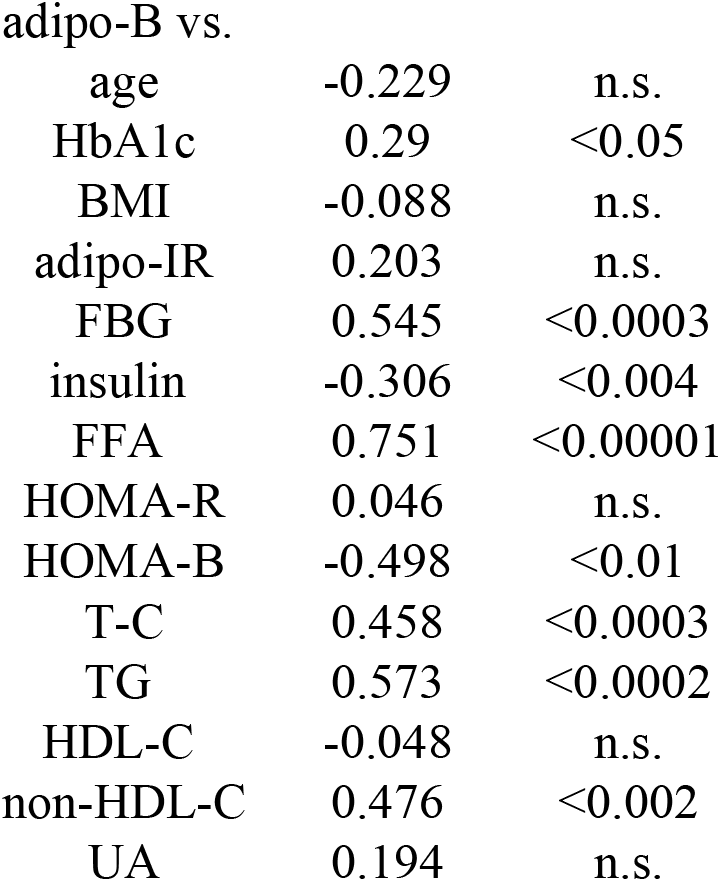
Correlation between adipo-B and other metabolic parameters. Simple regression analysis was performed between adipo-B and the indicated metabolic parameters.

### Factors Independently Associated with Lipid-Related Metabolic Parameters

To determine whether adipo-B was independently associated with lipid-related metabolic abnormalities, multivariable linear regression analyses were performed. As shown in Table 3A, 3B and 3C, adipo-B was independently associated with non-HDL-C. In addition, both adipo-B and HOMA-R were independently associated with FFA (Table 3A) and TG (Table 3B). These findings suggest that adipo-B is linked to lipid-associated metabolic abnormalities independent of conventional glycemic and anthropometric parameters.

**Table 3.**
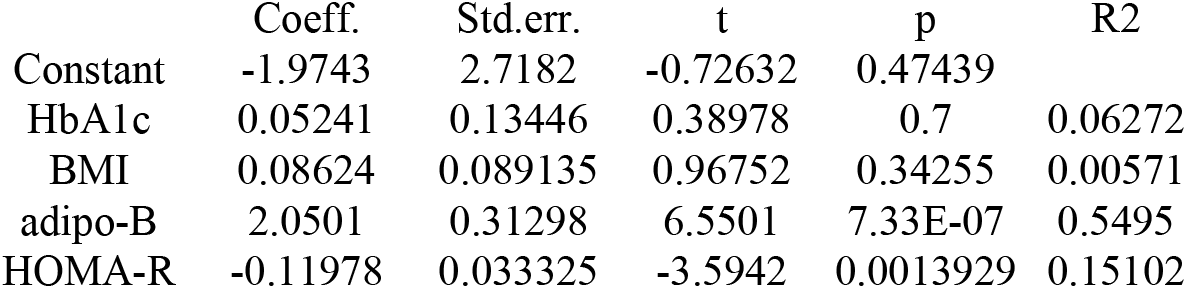
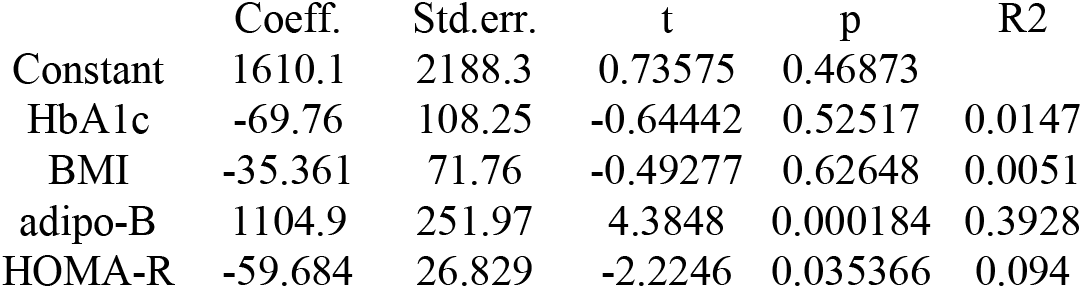
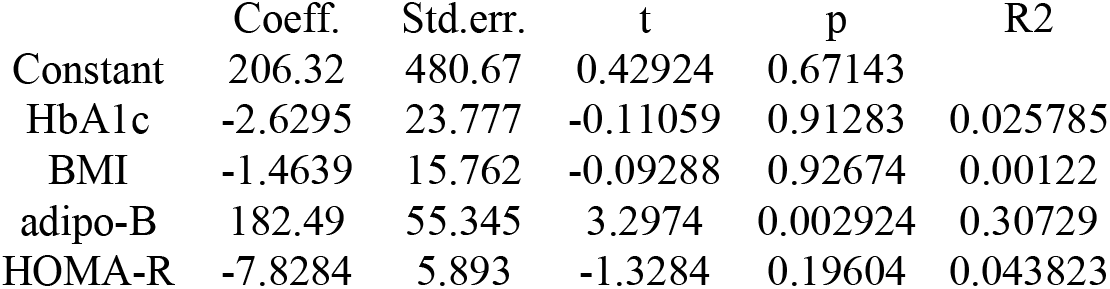
Factors independently associated with lipid-related metabolic parameters. Multivariable regression analysis was performed using the indicated parameters as the dependent variable, and HbA1c, BMI, adipo-B, and HOMA-R as independent variables. A) FFA B) TG C) non-HDL-C

### Metabolic Heterogeneity Despite Similar HbA1c and BMI

To further evaluate metabolic heterogeneity within clinically similar subjects, the subgroup was stratified according to the median adipo-B value into low adipo-B (Group A) and high adipo-B (Group B) groups. As shown in Table 4, subjects with high adipo-B exhibited significantly higher levels of FBG, FFA, TG, and non-HDL-C compared with subjects with low adipo-B, despite comparable HbA1c and BMI values. In addition, HOMA-B levels were significantly lower in the high adipo-B group. These findings indicate that subjects with apparently similar glycemic and anthropometric profiles may nevertheless exhibit substantially different underlying metabolic phenotypes.

**Table 4.**
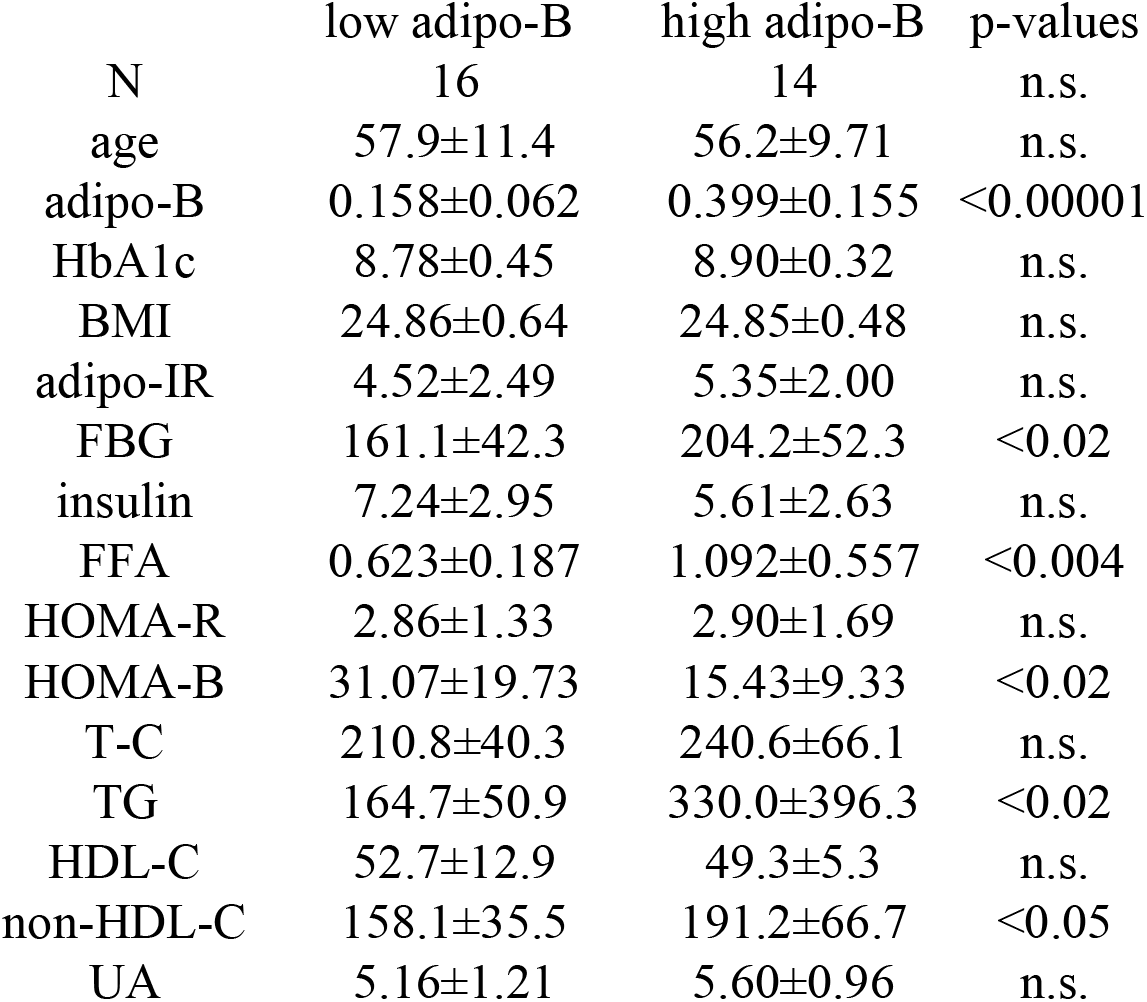
Baseline adipo-B dependent levels of metabolic parameters. The subjects were separated into two groups based on the median adipo-B value and unpaired Student t-test was performed between the two groups for the indicated metabolic parameters. The results are expressed as the mean±standard deviation (SD).

## Discussion

The newly diagnosed, treatment-naïve Japanese subjects with T2DM in this cohort exhibited poor glycemic control, modest overweight, and reduced insulin secretion and beta-cell function, along with relatively elevated TG and non-HDL cholesterol levels. HDL-C and UA remained within the normal range (Table 1). These features are consistent with previously reported findings in similar populations (4, 5).

The most striking finding in this present study is that such Japanese subjects exhibit substantial metabolic heterogeneity despite relatively similar HbA1c and BMI values. In particular, adipo-B identified distinct lipid-associated metabolic phenotypes within a clinically homogeneous subgroup characterized by similar glycemic severity and body size. In detail, while significant associations between adipo-B and glycemic parameters (FBG and HbA1c) and multiple lipid-related metabolic parameters including TG, non-HDL-C, and FFA were observed (Table 2), multivariable linear regression analysis demonstrated that these associations persisted after adjustment for potential confounding factors (Table 3A, 3B, 3C). Of note, HOMA-R showed no significant correlation with FFA or TG in simple regression analysis (Table 2), yet emerged as an independent predictor of both parameters in multivariable analysis (Tables 3A and 3B). This apparent discrepancy likely reflects suppression effects inherent to multivariable modeling. In simple regression, the association between HOMA-R and lipid parameters may be obscured by its collinearity with adipo-B, since both indices share insulin as a component variable. When adipo-B is included in the model and its variance is statistically accounted for, the independent contribution of HOMA-R to FFA and TG variability becomes apparent. This finding suggests that whole-body insulin resistance (as captured by HOMA-R) and the adipose-pancreas stress axis (as captured by adipo-B) contribute to lipid dysregulation through partially distinct mechanisms.

These findings suggest that adipo-B may capture aspects of metabolic dysfunction not adequately reflected by conventional glycemic or anthropometric markers alone. The subgroup analysis may be particularly informative. Even among subjects with similar HbA1c and BMI values corresponding to the most representative phenotype of newly diagnosed Japanese T2DM (Fig. 1A and 1B), marked differences in lipid-related metabolic parameters were observed according to adipo-B status. Subjects with high adipo-B exhibited higher FFA, TG, and non-HDL-C levels despite comparable HbA1c and BMI (Table 4). These observations suggest the presence of hidden metabolic heterogeneity that may not be readily detectable using conventional clinical markers. Notably, while no differences were noted with HbA1c, significantly higher FBG was observed in the high adipo-B group versus low adipo-B group (Table 4), suggesting that postprandial glucose regulation may also be linked to adipo-B.

The physiological interpretation of adipo-B remains speculative. Because adipo-B integrates adipo-IR and HOMA-B (7), this composite index may reflect the balance between adipose tissue metabolic stress and beta-cell compensatory capacity. Higher adipo-B values may therefore indicate disproportionate lipid-related metabolic stress relative to beta-cell function. However, the mathematical coupling among these indices should be carefully considered when interpreting the present findings.

Several limitations should be acknowledged. First, this study was cross-sectional and therefore causal relationships cannot be established. Second, the subgroup analysis involved a small number of subjects. Third, adipo-B is a mathematically derived composite index whose component variables overlap with some of the outcome measures examined, which should be considered when interpreting the observed associations. Finally, direct measurements of visceral adiposity, clamp-derived insulin sensitivity, or cardiovascular outcomes were not available.

## Conclusion

Newly diagnosed Japanese subjects with T2DM may exhibit substantial metabolic heterogeneity despite similar HbA1c and BMI values. Adipo-B may help identify unfavorable lipid-associated metabolic phenotypes not fully captured by conventional glycemic and anthropometric markers.

## Data Availability

All data produced in the present study are available upon reasonable request to the authors

## Abbreviations

T2DM: type 2 diabetes
BMI: body mass index
adipo-IR: adipose tissue insulin resistance
FBG: fasting blood glucose
FFA: free fatty acid
HOMA-R: homeostasis model assessment-R
HOMA-B: homeostasis model assessment-B
T-C: total cholesterol
TG: triglyceride
HDL-C: high density lipoprotein-cholesterol
UA: uric acid

